# Association of COVID-19 Infections in San Francisco in Early March 2020 with Travel to New York and Europe

**DOI:** 10.1101/2020.04.04.20053629

**Authors:** Wei Gu, Kevin Reyes, Elaine Hsu, Steve Miller, Charles Chiu

**Affiliations:** Department of Laboratory Medicine, University of California, San Francisco, California, USA; UCSF-Abbott Viral Diagnostics and Discovery Center, San Francisco, California, USA

## Abstract

Real-time dissemination of epidemiological survey data from positive COVID-19 cases is critical to support efforts to contain or reduce spread of viral infection in the community. Here we detected a significant association between domestic travel or travel to Europe and the identification of new cases in San Francisco, California, USA. These findings suggest that domestic and European travelers may need to be prioritized for evaluation of acute infection from COVID-19 in the setting of limited testing capacity.

---

As of April 4^th^, 2020, the COVID-19 pandemic caused by the novel SARS-CoV-2 coronavirus^1^ has infected more than 1.2 million people worldwide and the rise in cases has been exponential. In particular, New York cases in the United States surged from 22 to more than 113,000 between March 10 and now^2^, threatening to overwhelm hospitals and other regional health care systems in the city.

In San Francisco, we validated a qRT-PCR test to detect SARS-CoV-2 infection from nasopharyngeal swab samples based on the EUA (Emergency Use Authorization)- approved US CDC assay^3^. During the first 10 days since launch, we performed SARS-CoV-2 testing on 947 samples collected from March 10 through March 22 from patients with suspected SARS-CoV-2 infection at University of California, San Francisco. We analyzed the first 46 consecutive SARS-CoV-2 positive cases admitted to University of California, San Francisco hospitals or seen in outpatient clinics from March 10 through March 22. Data from these COVID-19 patients (**Table 1)** were matched with 102 randomly selected control patients over the same time period who tested negative for SARS-CoV-2. Documented history was recorded by a physician or nurse practitioner and included sick contacts, health care worker status, and travel history. Among the 46 COVID-19 patients, the median age was 44 years, 46% were female, and 65% were outpatients.

**Table 1.** Clinical Characteristics of COVID-19 Positive Cases (n=46)

|  |  |
| --- | --- |
| **Patient Demographics** |  |
| Age – years |  |
| Median (interquartile range) | 44 (33-62) |
| Range | (23-86) |
| Gender |  |
| Female – no. (%) | 21 (46%) |
| **Patient Characteristics** |  |
| Outpatient | 30 (65%) |
| Symptom Onset (median) | March 11, 2020 |
| Symptoms |  |
| Fever | 37 (80%) |
| Cough | 39 (85%) |
| Myalgias | 26 (57%) |
| Nasal congestion | 12 (26%) |
| Fatigue | 19 (41%) |
| Dyspnea | 14 (30%) |
| Diarrhea | 6 (13%) |
| White blood cell count ($10^9/L$) - median | 6.7 |
| Medical Risk Factors |  |
| Hypertension | 10 (22%) |
| Diabetes | 7 (15%) |
| Cardiac Disease | 5 (11%) |
| Lung Disease (asthma, COPD) | 3 (7%) |
| Immunocompromised (cancer, transplant) | 10 (22%) |

Abbreviations:
COPD - Chronic obstructive pulmonary disease

We noted that a travel history within 2 weeks of symptom onset (median date Mar 11, 2020) was significantly associated with COVID-19 infection (OR 3.8 [1.8-8.4]) **(Figure 1)**. Out of the 20 travelers with COVID-19 infection, there were significant associations for prior travel to Europe (5 travelers, OR 6.1 [1.1-32.7]), USA (14 travelers, OR 4.0 [1.6-10.0]), and specifically New York (6 travelers, OR 32.9 [1.8-598]) as compared to 17 travelers without COVID-19 infection **(Figure 2 and Table 2)**. No significant associations were found with regards to close contacts with known COVID-19 infected persons or frontline healthcare workers. Of note was one cluster of 3 positive cases associated with COVID-19 infection in an airport worker, that was categorized as a case of community rather than travel-associated transmission.

**Table 2.** Risk Factor Associations with COVID-19 Infection.

|  | Odds Ratio [CI 95%] | P value |
| --- | --- | --- |
| Healthcare Worker | 0.8974 [0.3442 to 2.3400] | P = 0.8248 |
| Close Contact | 2.8571 [0.9684 to 8.4295] | P = 0.0572 |
| Travel* | 3.8462 [1.7602 to 8.4043] | P = 0.0007 |
| NYC/New York* | 32.9012 [1.8114 to 597.5961] | P = 0.0182 |
| non-NY USA | 1.3800 [0.4695 to 4.0558] | P = 0.5582 |
| Europe* | 6.0976 [1.1368 to 32.7054] | P = 0.0349 |
| Asia | 2.2444 [0.1373 to 36.6894] | P = 0.5706 |

\* Significant associations at a p-value of 0.05 for significance

**Table 3.** Travel Locations by Patient History.

| **Infection Status** | **Geographic Location** | **Count** |
| --- | --- | --- |
| **COVID-19 Positive (n=29)** |  |  |
|  | New York metropolitan area or “New York” | 5 |
|  | Not New York, but USA | 9 |
|  | • Colorado | 2 |
|  | • Chicago | 1 |
|  | • Southern California (Los Angeles) | 1 |
|  | • Idaho | 1 |
|  | • Washington State | 1 |
|  | • Oregon | 1 |
|  | • Hawaii | 1 |
|  | • Washington DC | 1 |
|  | Europe | 5 |
|  | Asia | 1 |
| **COVID-19 Negative (n=25)** | New York metropolitan area or “New York” | 5 |
|  | Not New York, but USA | 9 |
|  | • Washington State | 4 |
|  | • Georgia | 2 |
|  | • Southern California | 2 |
|  | • Minnesota | 1 |
|  | • Texas | 1 |
|  | • Hawaii | 1 |

**Figure 1.**
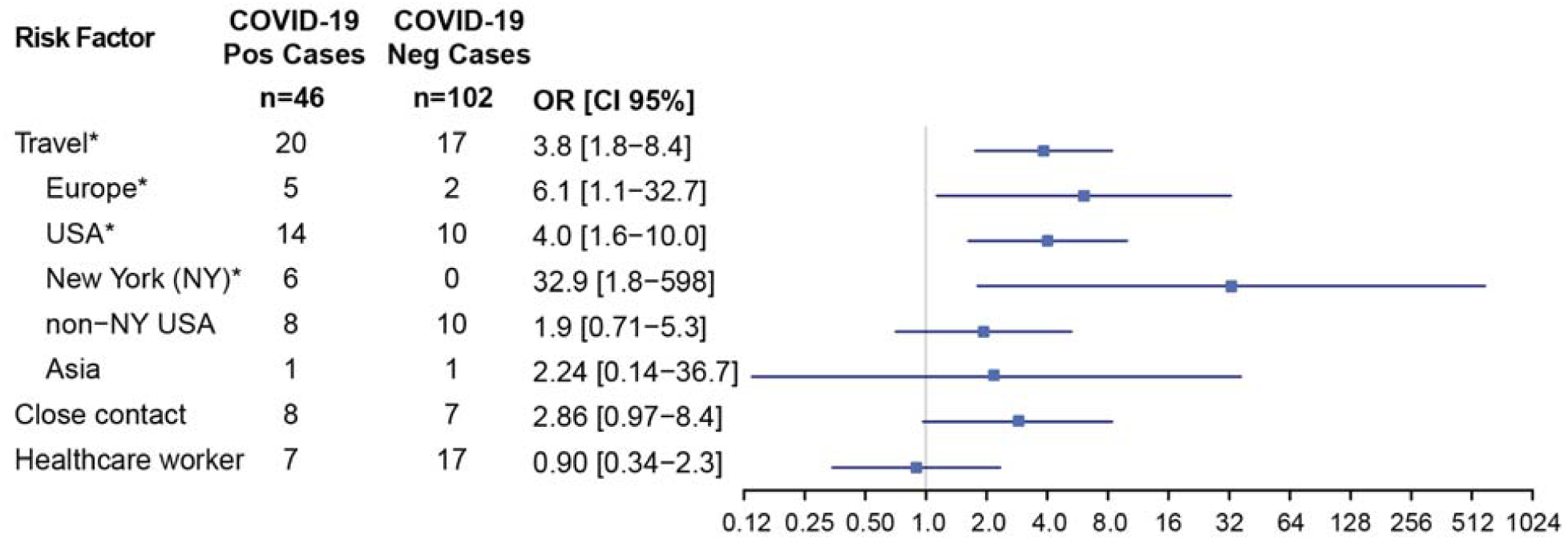
Associations with Positive COVID-19 RT-PCR Testing. The odds ratios (OR) with 95% confidence intervals (CI) are displayed. Positives (n=46) were consecutive cases from March 10 to 20, 2020. Negatives (n=102) were randomized from the same time period. Significant risk factors (p <0.05) are designated with ‘*’ and were recent travel, including Europe, USA (domestic), and/or New York. Abbreviations: Pos, positive; Neg, negative; OR, odds ratio; CI, confidence interval.

**Figure 2.**
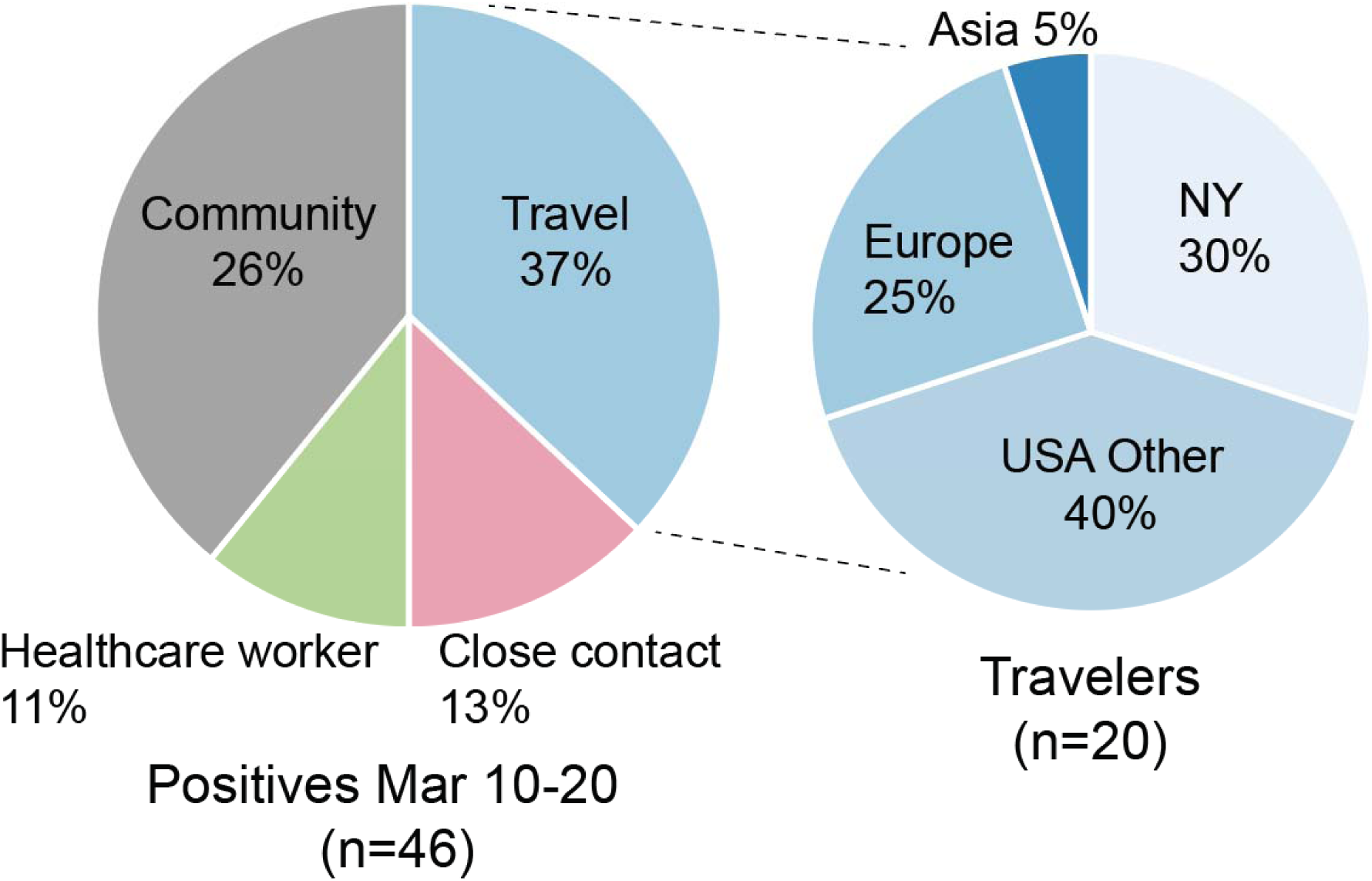
Travel History Associated with Positive Cases. All positive cases and their associations are shown here (left). Cases that overlapped in two categories (e.g. traveler and contact with a known positive, n=3) and were counted as a half-case in each category. Those who did not have recent travel history, close contact with a known COVID-19 positive, or a frontline healthcare worker, were uniformly categorized as a community case. The most common association with a positive case was a travel history immediately prior to symptoms. Travelers (n=20) are subdivided by travel region (right).

Our evaluation of diagnosed COVID-19 cases in San Francisco in early March 2020 associates with travelers from New York prior to the recognized spike in New York cases in late March **(Figure 3)**. Travel from New York was underrecognized as a risk factor for COVID-19 infection in the United States in early March. Guidelines for COVID-19 testing have not included screening for domestic travel. Our findings in San Francisco here can be extrapolated across America as there are over 100 direct domestic destinations and more than 6 million domestic flights a month from JFK, Newark, and LaGuardia airports^4^. In the near future, serological testing would be needed for surveillance of the much larger population of individuals with mild illness or asymptomatic infection^5^.

**Figure 3.**
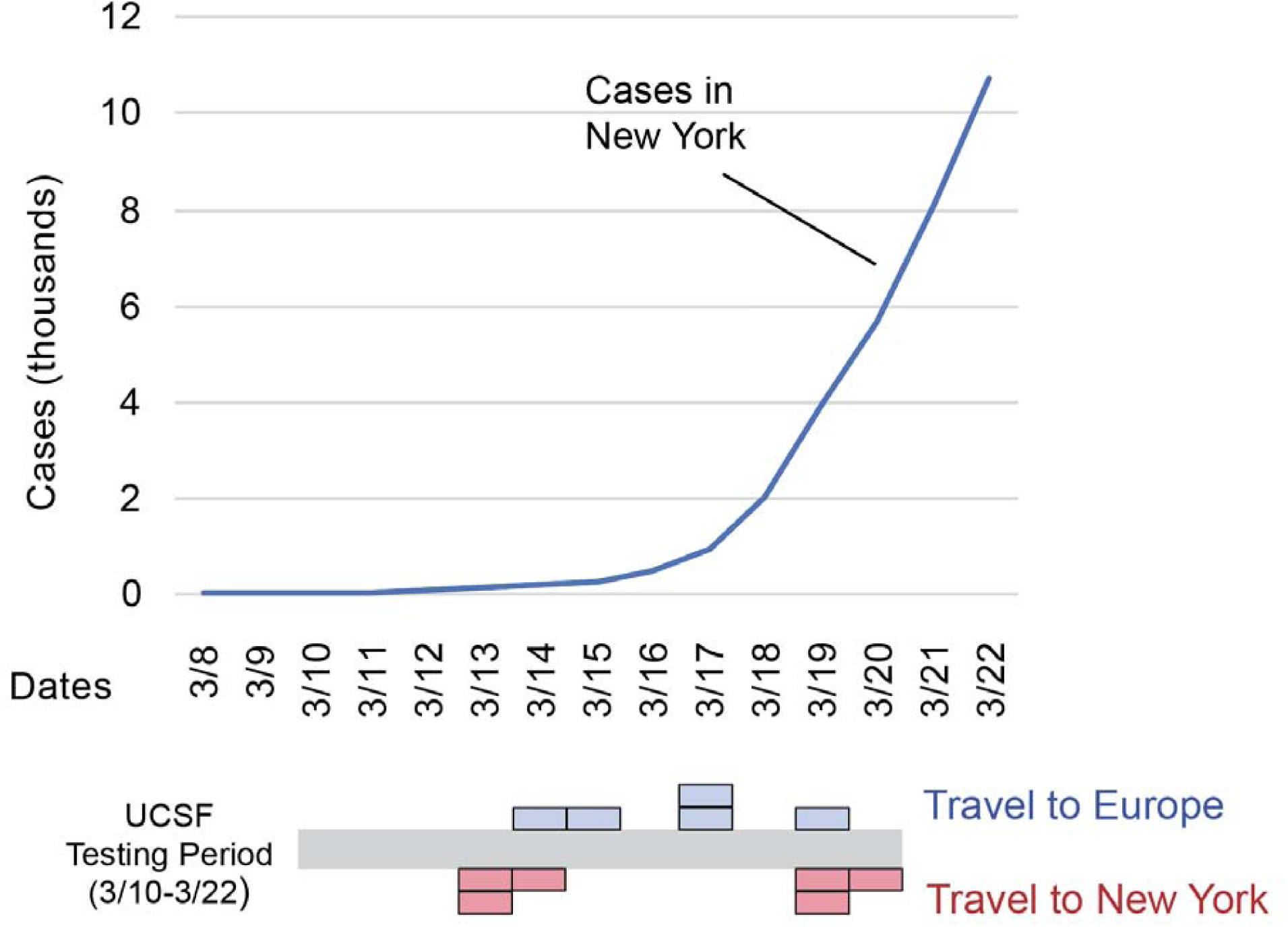
Timeline of cumulative COVID-19 cases diagnosed in New York, and UCSF positive cases found in San Francisco in association with New York or Europe over time. Each colored block represents a single patient.

## Data Availability

De-identified data from UCSF patients is available upon

## Ethics

The institutional review board at University of California, San Francisco approved this study. Non-identifying clinical, demographic, and laboratory data were extracted from the electronic medical board by retrospective chart review. Informed consent was waived for this minimal risk study.

## Data Availability

De-identified data from UCSF patients is available upon request.

